# Expert-level prenatal detection of complex congenital heart disease from screening ultrasound using deep learning

**DOI:** 10.1101/2020.06.22.20137786

**Authors:** Rima Arnaout, Lara Curran, Yili Zhao, Jami C. Levine, Erin Chinn, Anita J. Moon-Grady

## Abstract

Congenital heart disease (CHD) is the most common birth defect. Fetal survey ultrasound is recommended worldwide, including five views of the heart that together could detect 90% of complex CHD. In practice, however, sensitivity is as low as 30%. We hypothesized poor detection results from challenges in acquiring and interpreting diagnostic-quality cardiac views, and that deep learning could improve complex CHD detection. Using 107,823 images from 1,326 retrospective echocardiograms and surveys from 18-24 week fetuses, we trained an ensemble of neural networks to (i) identify recommended cardiac views and (ii) distinguish between normal hearts and complex CHD. Finally, (iii) we used segmentation models to calculate standard fetal cardiothoracic measurements. In a test set of 4,108 fetal surveys (0.9% CHD, >4.4 million images, about 400 times the size of the training dataset) the model achieved an AUC of 0.99, 95% sensitivity (95%CI, 84-99), 96% specificity (95%CI, 95-97), and 100% NPV in distinguishing normal from abnormal hearts. Sensitivity was comparable to clinicians’ task-for-task and remained robust on external and lower-quality images. The model’s decisions were based on clinically relevant features. Cardiac measurements correlated with reported measures for normal and abnormal hearts. Applied to guidelines-recommended imaging, ensemble learning models could significantly improve detection of fetal CHD and expand telehealth options for prenatal care at a time when the COVID-19 pandemic has further limited patient access to trained providers. This is the first use of deep learning to ∼double standard clinical performance on a critical and global diagnostic challenge.

## Introduction

Congenital heart disease (CHD), the most common birth defect ^1^, can be asymptomatic in fetal life but cause significant morbidity and mortality after birth ^1-3^. Compared to postnatal diagnosis, fetal diagnosis can improve neonatal outcomes, surgical/interventional planning^4-6^, and could enable *in utero* therapies^7,8^. Distinguishing normal fetal hearts from complex CHD requiring referral to a fetal cardiologist is therefore a critical and universal need, especially at a time when the COVID-19 pandemic has further limited patient access to prenatal care^9^. Low sensitivity in this task can limit palliation options, worsen postnatal outcomes, and hamper research on *in utero* therapies, while low specificity can cause unnecessary additional testing and referrals.

A fetal survey (fetal screening ultrasound) is recommended for every pregnancy worldwide ^10,11^ in the second trimester and generally includes five clinically recommended cardiac views (**Figure 1a**) that together could allow clinicians to diagnose up to 90% of complex CHD^12,13^. In practice, however, detection is often as low as 30%^1,14,15^, even where ultrasound is universal^10,11,15^. Specificity is also sub-optimal, as low as 40-50%^1^.

**Figure 1.**
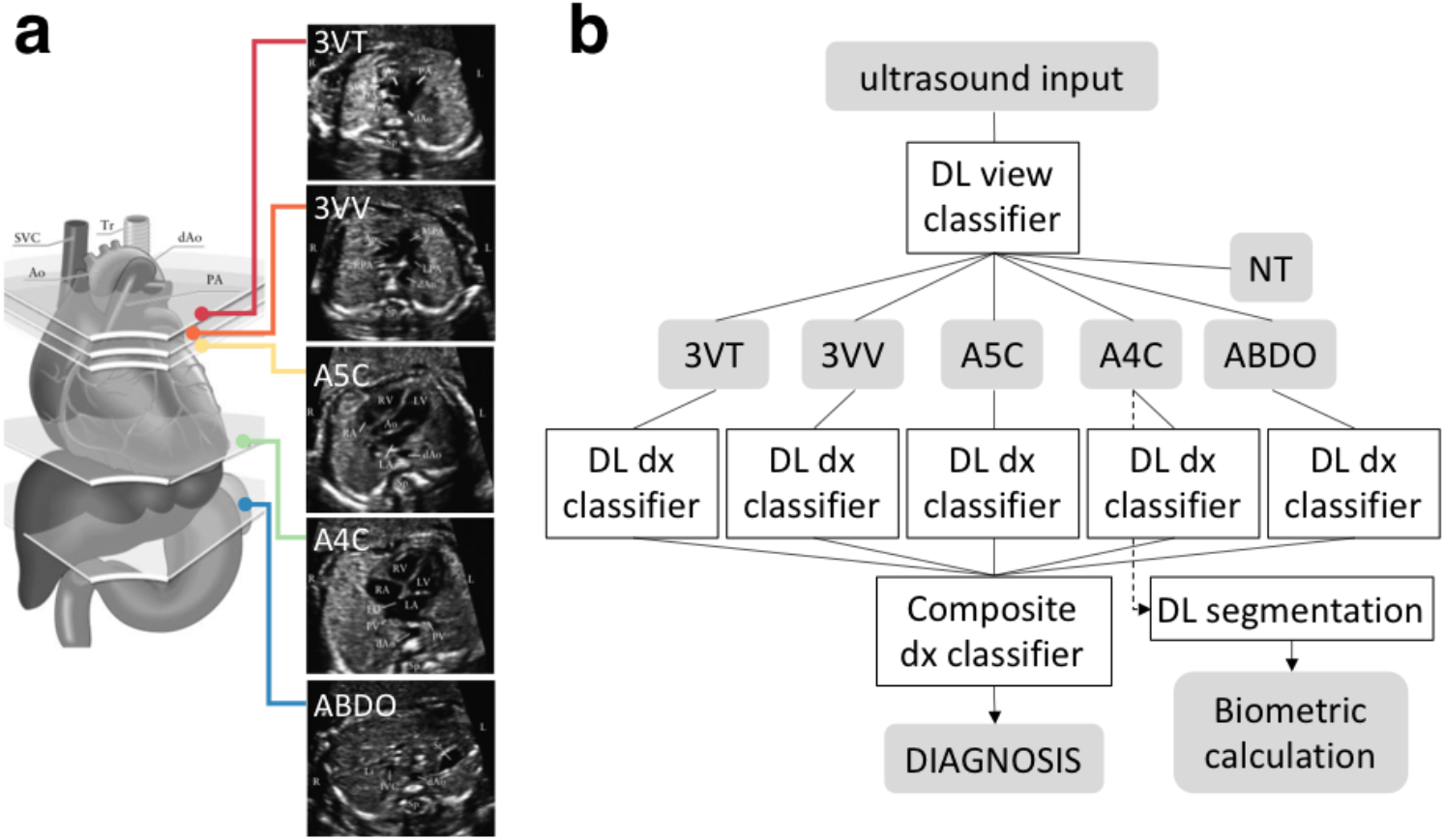
Overview of the ensemble model. (a) Guidelines recommend that the following five axial views can detect CHD. Illustration adapted with permission from Yagel et. al., Ultrasound Obstet Gynecol 2001;17(5):367-9. (b) The overall model is an ensemble of the components shown. From a fetal ultrasound, a deep learning classifier detects the five screening views (“DL view classifier”). Subsequent DL classifiers for each view detect whether that view is normal or abnormal (“DL dx classifiers”). These per-image, per-view classifications are fed into a rules-based classifier to create a composite diagnostic decision as to whether the fetal heart is normal or abnormal (“Composite dx classifier”). A4C views are also passed to a segmentation model to extract fetal cardiac biometrics. 3VT, 3-vessel trachea. 3VV, 3-vessel view. A5C, apical 5-chamber. A4C, apical 4-chamber. ABDO, abdomen. NT, non-target. DL, deep learning. dx, diagnosis.

Two reasons for this gap between possible and commonplace CHD detection are (i) inadequate expertise in interpretation and/or (ii) inadequate acquisition of diagnostic-quality images^16,17^. Causes of inadequate imaging include poor acoustic windows, fetal motion, and the small size of the fetal heart. Furthermore, a fetal survey includes thousands of image frames spanning multiple structures per single video “sweep,” so the diagnostic frames of interest for CHD may be just a handful and thus easily missed. Finally, the prevalence of CHD (∼0.8-1%) is low enough that non-experts see it only rarely and may discount or overlook abnormal images. Together, these make CHD detection one of the most difficult diagnostic challenges in ultrasound. Quality control studies aimed at addressing these challenges succeeded in increasing CHD detection considerably^18,19^, showing that improving image acquisition and interpretation can bridge the diagnosis gap; however, these small, single-center programs are difficult to sustain and scale.

Deep learning (DL) is a state-of-the-art type of machine learning useful in image analysis^20-23^. DL has been applied to adult cardiac ultrasound^24,25^, besting clinicians on view classification on small, downsampled datasets^26^. DL can be used to classify images or to segment structures within images; several DL models can be used together in an ensemble fashion. While hardly the only diagnostic challenge in ultrasound, fetal CHD detection is certainly one of the most challenging and is therefore a good use case for deep learning. We hypothesized that DL could improve ultrasound analysis for CHD.

## Methods

### Data sets

All datasets were obtained and de-identified, with waived consent in compliance with the Institutional Review Board (IRB) at the University of California, San Francisco (UCSF) and the IRB at Boston Children’s Hospital.

#### Inclusion, exclusion, and definitions of normal and CHD

Fetal echocardiograms and fetal surveys (second-trimester obstetric anatomy scans performed by sonographers, radiologists and/or maternal-fetal-medicine physicians) performed between 2000 and 2019 were utilized. Images came from GE (67%), Siemens (27%), Philips (5%), and Hitachi (<1%) ultrasound machines. Inclusion criteria were fetuses of 18-24 weeks of gestational age. Presence of significant non-cardiac malformations (e.g. congenital diaphragmatic hernia, congenital airway malformation, congenital cystic adenomatoid malformation, meningomyelocele) were excluded. Gold-standard definitions of normal vs. CHD were made as follows. CHD pathology was determined by review of the clinical report as well as visual verification of the CHD lesion for each ultrasound by clinician experts (Drs. Grady, Levine, and Zhao with over 60 years combined experience in fetal cardiology). Additionally, for studies performed in or after 2012, we were able to validate the presence, absence, and type of cardiac findings in the ultrasound studies with electronic health record codes for CHD in the resulting neonates (ICD-9 codes 745*, 746*, and 747* and ICD-10 codes Q2*, and ICD procedure codes 02*, 35*, 36*, 37* 38*, 88*, and 89*). Studies where clinician experts did not agree on the lesion and no post-natal diagnosis was present, were not included. Normal fetal hearts were defined as negative for structural heart disease, fetal arrhythmia, maternal diabetes, maternal lupus, maternal Sjögren syndrome, or presence or history of abnormal nuchal translucency measurement, non-cardiac congenital malformations, or CHD as defined above. Abnormal fetal studies had the presence of any of the following lesions: tetralogy of Fallot, pulmonary atresia with VSD, or double outlet right ventricle with VSD committed to aorta (TOF); critical aortic stenosis and hypoplastic left heart syndrome (HLHS); isolated aortic stenosis (AS); atrioventricular septal defect (AVSD); D-transposition of the great arteries (DTGA); L-transposition of the great arteries (LTGA); single ventricle, including heterotaxy with left or right atrial isomerism (SV); double outlet right ventricle with uncommitted, doubly committed, or sub-pulmonary VSD (DORV); aortic coarctation (COARCT); total anomalous pulmonary venous return (TAPVR); truncus arteriosus (TRUNCUS); Ebstein’s anomaly / tricuspid dysplasia (EB); tricuspid atresia (TA); and pulmonary atresia with intact IVS (PAIVS). Isolated VSD was not included as they only rarely require perinatal intervention.

#### Study design, training, and test sets

We analyzed images from a retrospective cohort. The total number of CHD echocardiograms, and the need to limit class imbalance between normal and CHD studies in training, were constraints guiding development of training and test datasets (**Figure S1d**). We first took all fetal echocardiograms with CHD fitting inclusion/exclusion criteria above (437 studies). To reduce class imbalance in training, we then took a sample of normal fetal echocardiograms (875 studies) such that CHD was ∼30 percent of the dataset. From this overall UCSF dataset, we created UCSF training and test sets as follows. We found those fetal echocardiograms which had a corresponding fetal survey in the UCSF system; a random sample of ∼10 percent from each lesion class made up FETAL-125 and OB-125, respectively (corresponding echocardiograms and surveys, respectively, from the same patients). FETAL-125 comprised 11,445 normal images and 8,377 abnormal images; OB-125 comprised 220,990 normal images and 108,415 abnormal images. The remaining ∼90 percent of fetal echocardiograms (1,187) were used for training, supplemented by 139 normal fetal surveys (1,326 studies total). For a population-based UCSF testing set, we started with OB-125 and added additional 3,983 normal fetal surveys such that the CHD lesions in OB-125 comprised 0.9% of an overall dataset totaling 4,108 surveys. The result was OB-4000, which comprised 4,473,852 images. As an external testing set, we received 423 fetal echocardiograms (4,389 images from 32 normal studies and 40,123 images from 391 abnormal studies) from Boston Children’s Hospital. These training and test sets are summarized in **Table 1** and **Figure S1d**.

**Table 1.**
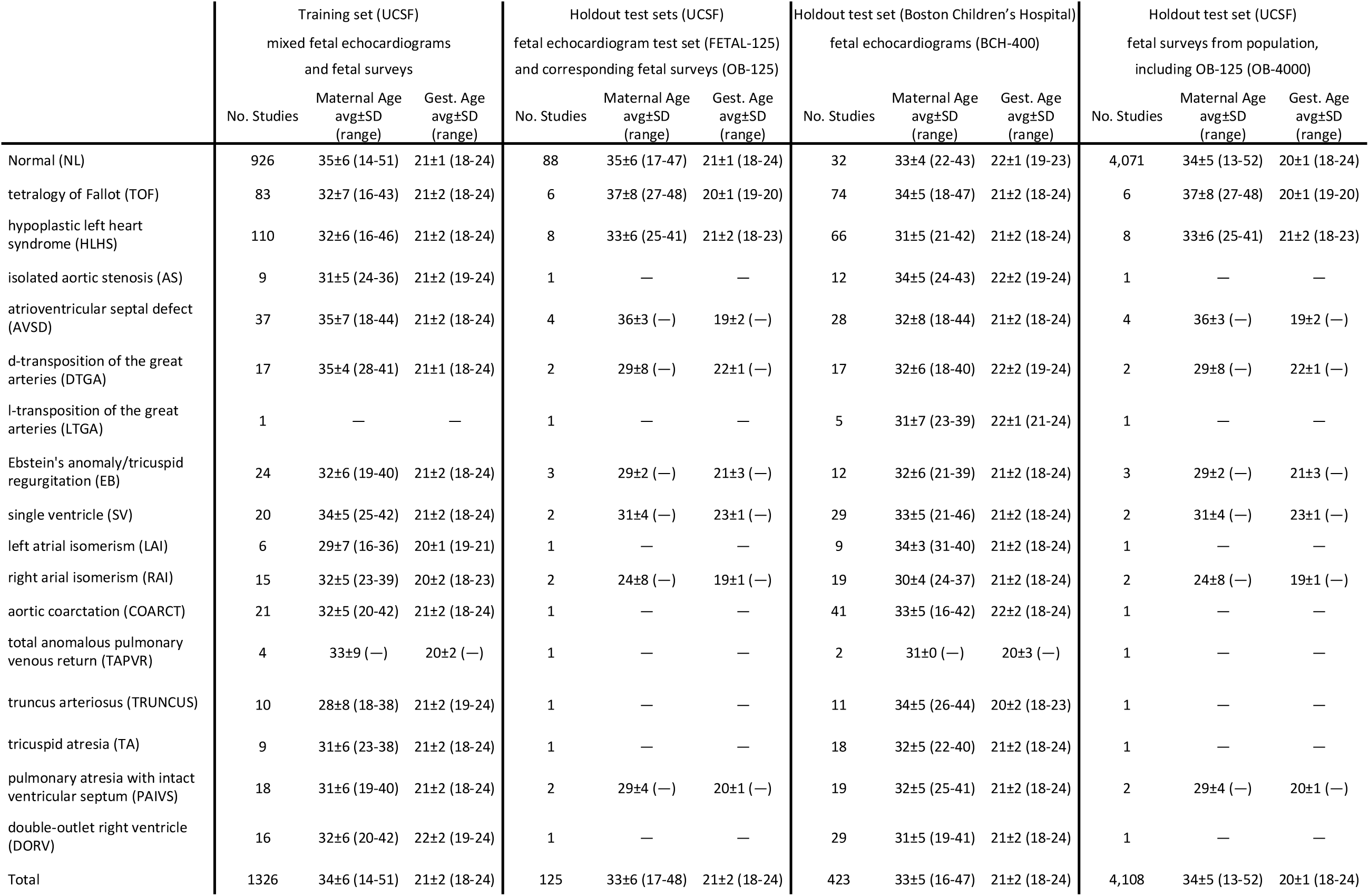
Demographics of Training and Test Sets. In small groups where there is a dash, information was withheld to protect patient privacy.

Separately, we obtained a test set of 10 twin ultrasounds between 18-24 weeks of gestational age (5,754 echocardiogram images, 36,355 fetal survey images). Eight sets of twins had normal hearts; one set of twins had one normal, one TOF heart; and one set of twins had one normal, one HLHS heart.

The above training dataset was used to train (i) a view classifier, (ii) normal vs. abnormal diagnostic classifiers for each target view, and (iii) a segmentation model. For the view classifier, 53,532 images from the 926 normal hearts were used. For the per-view diagnostic classifiers, 46,498 of the above images from 916 normal hearts were combined with an additional 54,291 images from 400 abnormal hearts (for a total of 1,316 studies, 100,789 images). (Ten of the studies used in training the view classifier only had non-target views and so were not used to train the diagnostic classifiers.) For segmentation of cardiac chambers, 1248 apical 4-chamber (A4C) images from 186 studies (122 normal, 25 HLHS, 39 TOF) were used. For segmentation of heart and thorax, 952 A4C images from 223 studies (157 normal, 25 HLHS, 41 TOF) were used. For all trainings, roughly equal proportions of data classes were used. Every image frame of the training set, FETAL-125, OB-125, and BCH-400 were view-labeled by clinician experts (approximately 20% of the dataset was independently scored by both labelers to ensure agreement). Because OB-4000 was too large for this approach, experts instead verified only that the top five predictions from the view classifier did in fact contain views of interest before that study underwent diagnostic classification. To maintain sample independence, training and test sets did not overlap by image, patient, or study.

### Data processing

DICOM-formatted images were deidentified as previously described^26^. Axial sweeps of the thorax were split into constituent frames at 300 by 400-pixel resolution. For view classification tasks, images were labeled as 3-vessel trachea (3VT), 3-vessel view (3VV), apical 5-chamber (A5C), apical 4-chamber (A4C), and abdomen (ABDO). A sixth category, called non-target (NT), comprised any fetal image that was not one of the five cardiac views of interest. For disease classification tasks, studies were labeled by normal or CHD lesions mentioned above.

For input into classification networks, each image was cropped to 240 x 240 pixels and downsampled to 80 x 80 pixels and scaled with respect to greyscale value (rescale intensity). For input into segmentation networks, images were cropped to 272 x 272 pixels and scaled with respect to greyscale value. All preprocessing steps made use of open-source Python libraries OpenCV (https://opencv.org/), Scikit-image (https://scikit-image.org/), SciPy (https://www.scipy.org/), and NumPy (https://numpy.org). For training fetal structural and functional measurements, OpenCV was used to label thorax, heart, right atrium, right ventricle, left atrium, left ventricle and spine from A4C images.

### Model Architecture and Training

#### Classification models

Classification models were based on the ResNet architecture^27^, with the following modifications. For view classification, batch size was 32 samples and training was over 175 epochs using the Adam optimizer and an adaptive learning rate (0.0005 for epochs 1-99; 0.0001 for epochs 100-149, and 0.00005 at 150+ epochs). Dropout of 50% was applied prior to the final fully-connected layer. Data were augmented at run-time by randomly applying rotations of up to 10 degrees, width and height shifts of up to 20 percent of total length, zooms of up to 50 percent, and vertical/horizontal flips. For diagnostic classification, transfer learning was applied to the previously described view classification model as follows: the first 18 layers were frozen. Additional training used the above settings except epochs ranged from 12 to 60, learning rate was constant for each model, no adaptive learning was used, and learning rate ranged from 0.00001 to 0.0001. Loss function was categorical cross-entropy (view classifier) or binary cross-entropy (diagnostic classifiers). Classification network architecture is shown in **Figure S1a**. Training and validation datasets in which view labels were randomized were used as a negative control, resulting in an F-score commensurate with random chance among classes.

#### Segmentation model

A4C images with clinician-labeled cardiothoracic structures (thorax, heart, spine, and each of the four cardiac chambers) were used as training inputs to a U-Net^28^ neural network architecture with modifications as in **Figure S1b**. Two different models were trained to detect (i) heart, spine, and thorax, and (ii) the four cardiac chambers. Batch size was 2, models were trained for 300-500 epochs, and an Adam optimizer was used with adaptive learning rates of 0.0001 to 0.00001. For data augmentation, width/shift was set at 20 percent, zoom was 15 percent, random rotations of up to 25 degrees, and horizontal/vertical flips were used. Loss function was categorical cross-entropy.

#### Framework and training and prediction times

All models were implemented in Python using Keras^29^ (https://keras.io/) and a Tensorflow (https:www.tensorflow.org/) backend. Trainings were performed on Amazon’s EC2 platform with a GPU instance p2.xlarge and took about 1.95 – 5h for segmentation models and 6 minutes – 4.6h for classification models. Prediction times per image averaged 3 ms for classification and 50 ms for segmentation on a standard laptop (2.6 GHz Intel core, 16GB RAM).

#### Use of prediction probabilities in classification

For each classification decision on a given image, the model calculates a probability of the image belonging to each of the possible output classes; as a default, the image is automatically assigned to the class with the highest probability. In certain testing scenarios, a threshold of acceptable prediction probability was applied to view classifications: namely, for OB-4000 “high confidence” views, diagnostic classification was performed only on images with view prediction probabilities greater than the first quartile for each view, and for OB-125 “low-quality” views, views with a model-predicted probability ≥0.9, but that human labelers did not choose as diagnostic quality, were used (**Results, Table S1**). A probability threshold for diagnostic classifications was also used in the rules-based composite diagnostic classifier, described below.

#### Quantification of cardiothoracic ratio, chamber fractional area change, and cardiac axis

Cardiothoracic ratio was measured as the ratio of the heart circumference to the thorax circumference. Fractional area change for each of the four cardiac chambers was calculated as [maximum pixel area – minimum pixel area]/[maximum pixel area]. Cardiac axis was calculated as the angle between a line centered on the spine and thorax, and a line centered on either the left chambers or the right chambers, whichever side had the greatest area. (The line centered on the cardiac chambers was chosen as a computational method of finding a line parallel to the intraventricular septum, used clinically and in ground-truth labeling.) Various checks were implemented to prevent calculation of clinical values from images with poor segmentation results. Concordance of predicted quantitative measurements were compared to ground truth measures (labeled images, and clinical measurements where available) using the Mann-Whitney U test. Measurements among normal, TOF, and HLHS groups were compared using the Kruskal-Wallis test.

#### Composite diagnostic classification

A rules-based classifier (“Composite dx classifier,” **Figure 1b**) was developed to unite per-view, per-image predictions into a single composite decision of normal vs. CHD. (We chose a rules-based approach because a machine learning-based classifier at this step would have required sacrificing some of our test data to train the composite classifier.) The rules-based composite diagnostic classifier operates as follows (**Figure S1c**). Only views with AUC > 0.85 (**Figure 3a**) were used. For each of the cardiac views of interest, a variable number of images each held a probability *p*_*CHD*_ of CHD for each image; the probability of normal for that image was also recorded, where *p*_*NL*_ = 1-*p*_*CHD*_. *p*_*CHD*_ below a certain cutoff threshold determined by the ROC curve for each view (**Figure 3a**) were reset to 0 to avoid over-scoring CHD. The resulting *p*_*NL*_’s and *p*_*CHD*_’s for each view were then summed separately, in order to maintain a distinction between a view being present-and-normal vs. being missing from a study, and each was normalized by the total sum of all predictions to account for different numbers of images in each view class to obtain the view-specific prediction values:

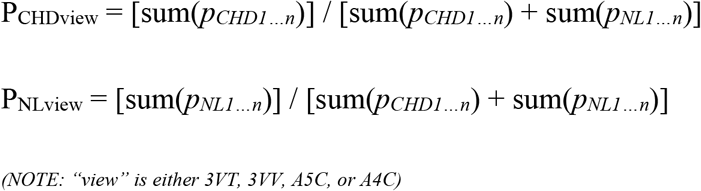

These prediction values for each view were in turn summed for a composite classification. Evaluating true positive, false positive, true negative, and false negative with different values for the threshold number allowed construction of a ROC curve (**Figure 3e**).

### Model Evaluation

Overall accuracy, per-class accuracy, average accuracy, confusion matrices, F-scores, receiver operator characteristics, C-statistics, and saliency maps (guided backpropagation) were calculated as previously described^26,30^. GradCAM was also used as previously described^31^. For performance analysis of segmentation models, Jaccard similarities were calculated in the standard fashion as the intersection of predicted and labeled structures divided by their union.

### Comparison to Human Experts

Clinicians with expertise in fetal cardiology (fetal cardiology and maternal-fetal medicine attendings, experienced fetal cardiology sonographers, fetal cardiology fellows, n=7), were shown up to one image per view for the studies in the OB-125 test set and asked whether that study was normal or not. For segmentation, clinical labelers segmented a subset of images multiple times, and intra-labeler Jaccard similarities were calculated as a benchmark. Use of clinicians for validation was deemed exempt research by the UCSF CHR.

### Data and Code Availability

Due to the sensitive nature of patient data (and especially fetuses as a vulnerable population), we are not able to make these data publicly available at this time. ResNet and UNet are publicly available (e.g. at https://keras.io/examples/cifar10_resnet/ and https://github.com/zizhaozhang/unet-tensorflow-keras/blob/master/model.py) and can be used with the settings described above and in **Figure S1**. Additional code will be available upon peer-reviewed publication at https://github.com/ArnaoutLabUCSF/cardioML

## Results

To test whether DL can improve fetal CHD detection, utilizing multi-modal imaging and experts in fetal cardiology, we implemented an ensemble of neural networks (**Figure 1b**) to (i) identify five diagnostic-quality, guidelines-recommended cardiac views (**Figure 1a**) from among all images in a fetal ultrasound (survey or echocardiogram), (ii) use these views to provide classification of normal vs. any of 16 complex CHD lesions (**Table 1**), and (iii) calculate cardiothoracic ratio (CTR), cardiac axis (CA), and fractional area change (FAC) for each cardiac chamber.

To train the various components in the ensemble, up to 107,823 images from 1,326 studies were used (**Table 1**). Several independent test datasets were used for evaluating model performance: 125 UCSF fetal echocardiograms (FETAL-125, 19,822 images) and 125 corresponding fetal surveys from the same patients (OB-125, 329,405 images), each with 30% CHD; a population-based sample of 4,108 fetal surveys with 0.9% CHD (4,473,852 images; includes OB-125), and an external set from Boston Children’s Hospital consisting of 423 fetal echocardiograms highly enriched for CHD (BCH-400 with 44,512 images, 92% CHD) (**Table 1, Methods, Figure S1d**). Prediction times per image averaged 3 milliseconds for classification and 50 milliseconds for segmentation on a standard laptop (**Methods**).

### View classification

Identifying the five views of the heart recommended in fetal CHD screening^12^—3-vessel-trachea (3VT), 3-vessel view (3VV), apical-5-chamber (A5C), apical-4-chamber (A4C), and abdomen (ABDO)—was a prerequisite for diagnosis. We therefore trained a convolutional neural network^27^ (**Figure S1a**) view classifier (“DL view classifier”, **Figure 1b**) to pick the five screening views from fetal ultrasound, where any image that was not one of the five guidelines-recommended views was classified as “non-target” (NT; e.g. head, foot, placenta). Training data was multi-modal including both fetal echocardiograms, which naturally contain more and higher-quality views of the heart, and fetal surveys, offering a full range of non-target images. Notably, only views of sufficient quality to be used for diagnosis (as deemed by expert labelers, see **Methods**) were used to train the view classifier.

On normal studies in the FETAL-125 test set, the F-score (the harmonic mean of precision and recall) for view classification was 0.93, (AUC range 0.94-0.98, **Figure 2a, 2b**). The network’s classification decision on a particular image is determined by the probability of the image belonging to each of the possible classes; by default, the image is assigned to the class with the highest probability. For fetal view classification, as demonstrated for adults^26^, mean probability for correct predictions was significantly higher than for incorrect (p-value Mann-Whitney U test, <1^e-300^)(**Figure 2c**).

**Figure 2.**
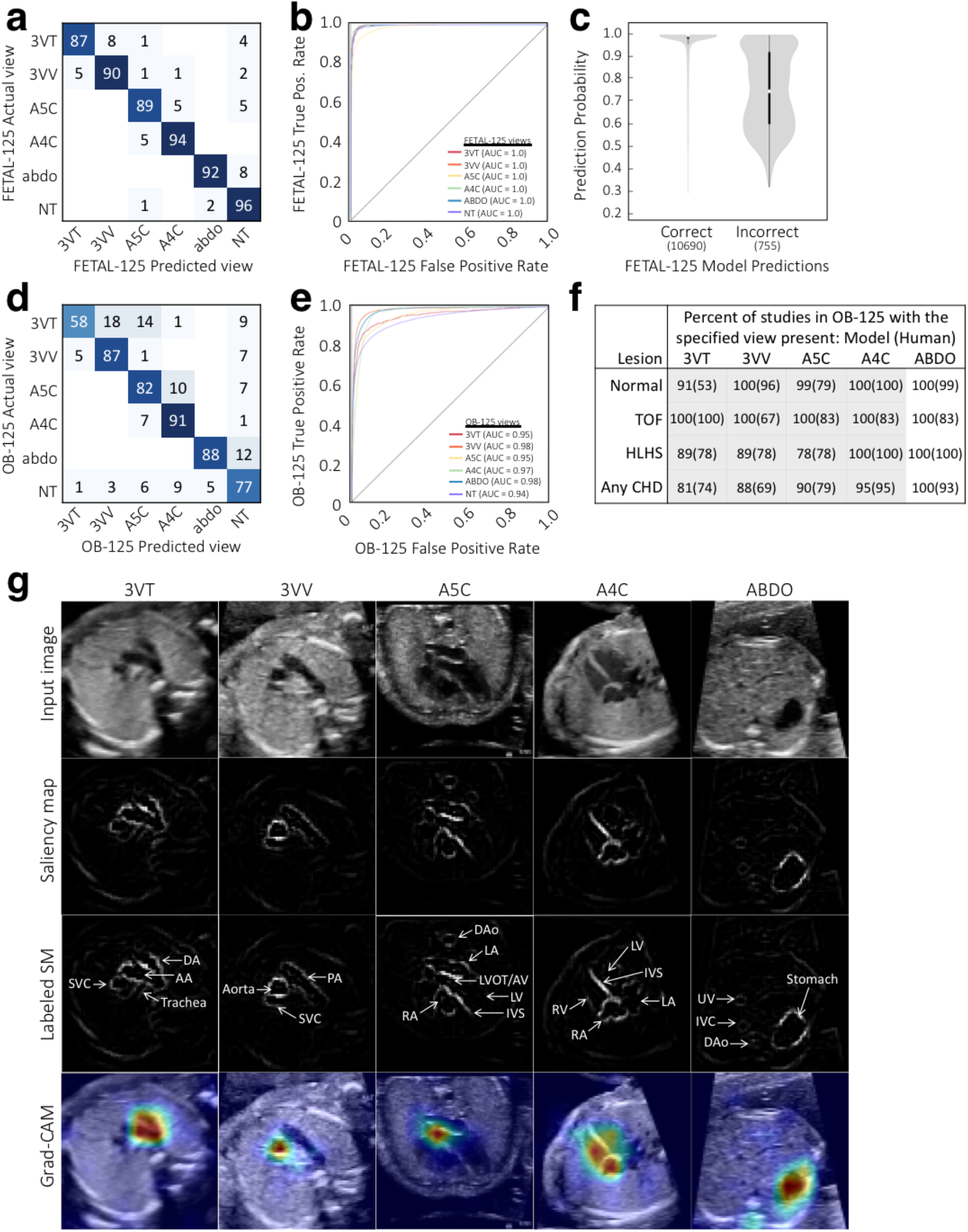
View detection performance. Normalized confusion matrix (a) and ROC curve (b) showing classifier performance on normal hearts from FETAL-125 test set. (c) Violin plots showing prediction probabilities for this test data show that when predictions are correct, predicted classification probability is high, while probability ranged for incorrect predictions. In violin plots, white dots signify mean, thick black line signifies 1^st^ to 3^rd^ quartiles. Normalized confusion matrix (d) and ROC curve (e) showing classifier performance on the OB-125 test set. (f) Percent of fetal surveys from the OB-125 test set with model-detected views (compared to human-detected views in parentheses). Grey shading indicates views with AUC ≥ 75 for normal/abnormal prediction from Fig. 3a,3d. (g) One example test image shown per view (top row), with corresponding saliency map (unlabeled, second row; labeled, third row) showing that the pixels most important to the model in predicting the view highlighted anatomic structures important to each view. Fourth row: gradient-weighted class activation map (Grad-CAM) provides a heatmap of regions of the image most important to the model in predicting the view. Grad-CAMs were also highly specific for structures distinguishing each view; the confluence of the aortic and ductal arches compared to the aortic cross-section distinguishing 3VT from 3VV, for example, and the left ventricular outflow tract vs. right heart distinguishing A5C from A4C. SM, saliency map, 3VT, 3-vessel trachea. 3VV, 3-vessel view. A5C, apical 5-chamber. A4C, apical 4-chamber. ABDO, abdomen. DA, ductal arch. AA, aortic arch. SVC, superior vena cava. PA, pulmonary artery. TV, tricuspid valve. AV, aortic valve. MV, mitral valve. IVS, interventricular septum. IAS, interatrial septum/foramen ovale.

We then tested the view classifier on OB-125 (**Figure 2d, 2e**). When diagnostic-quality target views were present, the view classifier found them with 90% sensitivity (95%CI, 90%) and 78% specificity (95%CI, 77-78%). Using only images with prediction probabilities at or above the first quartile, sensitivity and specificity increased to 96% and 92% (95%CI, 96% and 92-93%). Recommended views were not always present in each fetal survey and were more commonly present in normal studies (**Figure 2f**). The view classifier’s greatest confusion was between 3VT and 3VV (**Figure 2d**), adjacent views that often cause clinical uncertainty also^12,17,32^.

To validate that the view classifier utilized clinically relevant features, we performed both saliency mapping and gradient-weighted class activation mapping (Grad-CAM) experiments^26,31^ on test images to show the pixels (saliency mapping) or region (Grad-CAM) most important to the classifier in making its decision. Both experiments show that the view classifier makes its decisions based on clinically relevant image features (**Figure 2g**).

### Classification of normal vs. complex CHD

We trained the same convolutional neural network architecture used above to classify normal vs. CHD for each of the five view classes (“DL dx classifiers,” **Figure 1b**). On FETAL-125, AUC ranged from 0.72 (ABDO) to 0.88 (3VV and A4C; **Figure 3a**). Across all test datasets, AUCs for ABDO view reflected the clinical finding that abdomen is the least useful for CHD diagnosis. For each heart, we arrived at a composite diagnostic decision of normal vs. CHD by applying a rules-based classifier (“Composite dx classifier” **Figure 1b)** to the per-image, per-view predictions (**Methods, Figure S1c**).

**Figure 3.**
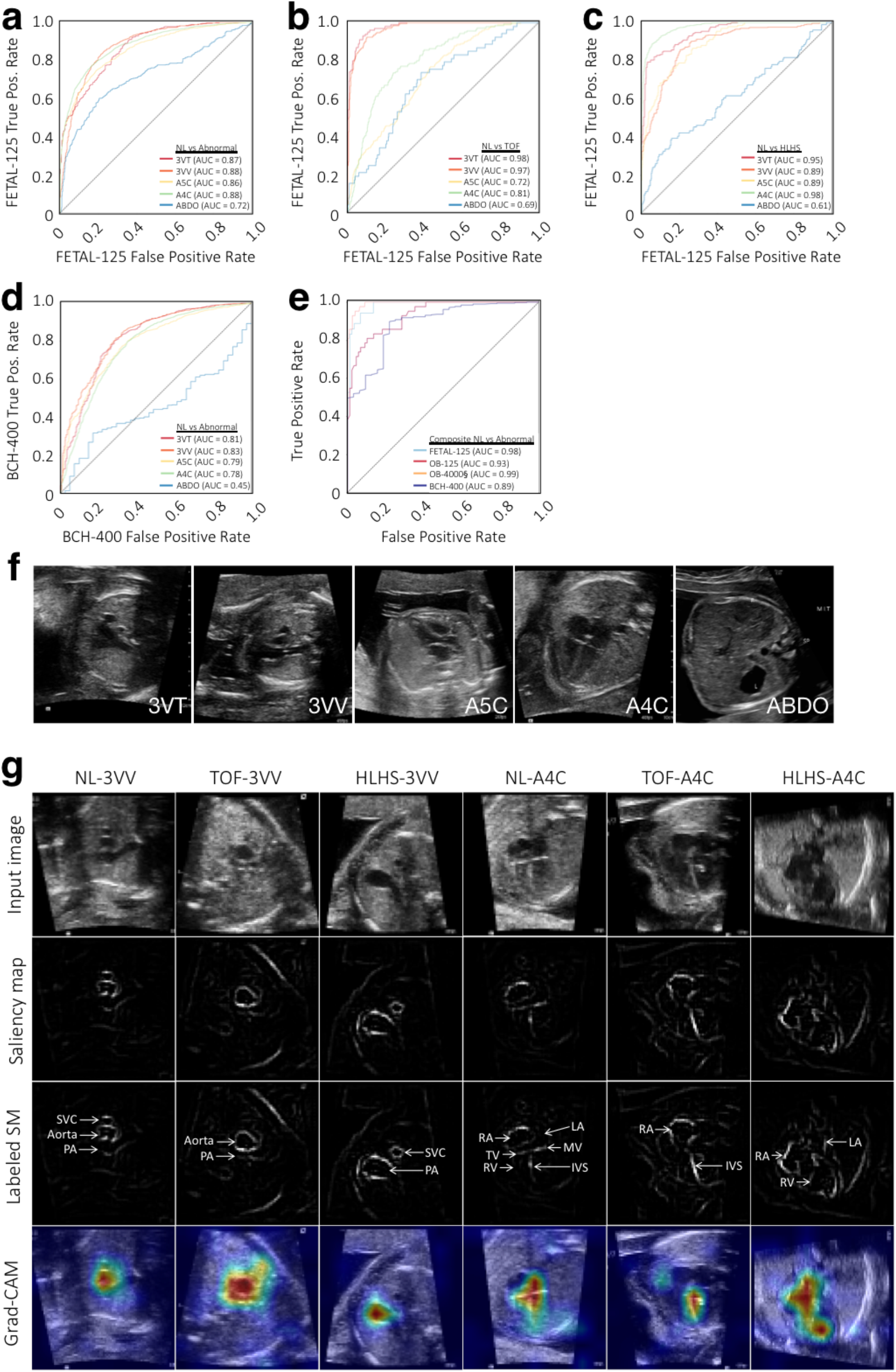
Performance of diagnostic classification. ROC curves showing model’s ability to distinguish (a) normal vs. any CHD lesion in Table 1, (b) normal vs. tetralogy of Fallot (TOF), and (c) normal vs. hypoplastic left heart syndrome (HLHS), for each of the five views in the FETAL-125 test dataset (OB-125 demonstrated similar findings, not shown). In (b) and (c), the views most clinically important for diagnosis of TOF and HLHS, respectively, are also those with the highest AUC. (d) ROC curve for prediction of per-view normal vs. abnormal from external data (BCH-400 test set). (e) ROC curve for composite (per-heart) prediction of normal vs. abnormal for each of the test datasets. “OB-4000§” indicates the high-confidence target images from OB-4000 test set (images with view-prediction probability at or above the first quartile). (f) Example of images given to both the model and clinicians for determination of normal vs. abnormal in a head-to-head comparison. (g) Top row: one example test image shown for normal, tetralogy of Fallot (TOF), and hypoplastic left heart syndrome (HLHS); three-vessel view (3VV) and apical four chamber view (A4C) shown. Second row: corresponding unlabeled saliency map. Third row: labeled saliency map, both showing the pixels most important to the model in classifying normal vs. abnormal. Fourth row: gradient-weighted class activation map (Grad-CAM) provides a heatmap of regions of the image most important to the model in prediction. In 3VV the relative sizes of aorta and pulmonary artery distinguishes these lesions from normal, and in A4C the angled intraventricular septum and enlarged right heart distinguish TOF and HLHS, respectively, from normal. SM, saliency map, 3VT, 3-vessel trachea. 3VV, 3-vessel view. A5C, apical 5-chamber. A4C, apical 4-chamber. ABDO, abdomen. SVC, superior vena cava. PA, pulmonary artery. TV, tricuspid valve. RA, right atrium. TV, tricuspid valve. RV, right ventricle. LA, left atrium. MV, mitral valve. IVS, interventricular septum.

Using this approach, we achieved AUCs of 0.98, 0.93, 0.99, and 0.89 in distinguishing normal from abnormal hearts on FETAL-125, OB-125, OB-4000, and BCH-400, respectively (**Figure 3e**). (To achieve this AUC for OB-4000, only images with view-prediction probabilities above the first quartile were used in the composite diagnostic classifier.) This allowed a sensitivity of 95% (95%CI, 83-99%), specificity of 96% (95%CI, 95-97%), positive predictive value (PPV) of 20% (95%CI, 17-23%), and negative predictive value (NPV) of 100% in OB-4000. Performance on these and additional tests discussed below are summarized in **Table S1**. Overall, model sensitivity on fetal echocardiograms rivaled that cited across several papers^33-35^ (p-value 0.3, assuming normal distribution of measures in the literature). More importantly, model sensitivity and specificity on fetal surveys was significantly better than reported performance^1,14,15,34^ (p-values 0.002 and 0.04, respectively).

We also wished to compare model performance on fetal surveys (OB-125) directly against clinicians. Therefore, we gave each the following test: one, full-resolution image per view, only five images in total per heart (**Figure 3f**). This test was chosen both to make the task feasible for humans, and, given the potential regional variation in image acquisition protocols, to simulate a “lean protocol” in which only minimal recommended views are acquired. Thirty-eight of the 125 fetal surveys (30%) in OB-125 contained all five views. On this test, the model achieved 88% sensitivity (95% CI, 47-100%) and 90% specificity (95% CI, 73-98%). Clinicians (n=7) achieved an average sensitivity of 86% (95% CI, 82-90%) and specificity of 68% (95% CI, 64-72%). The model was comparable to clinicians (p=0.3) in sensitivity and superior (p=0.04) in specificity.

To validate that the model generalizes beyond the medical center where it was trained^36^, we tested it on fetal echocardiograms from an unaffiliated, geographically remote medical center (BCH-400; **Table 1**). AUCs for view detection ranged from 0.95-0.99 (not shown). AUC for composite classification of normal vs. abnormal hearts was 0.89, despite a high prevalence of abnormal hearts in this test set (**Figure 3e, Table S1**).

Multifetal pregnancies have a higher risk of CHD than the general population^1^. Therefore, a CHD detection model applicable to ultrasounds of twins and other multiples would be useful. Based on saliency mapping and Grad-CAM experiments (**Figures 2g, 3g**), we hypothesized our model could perform adequately on surveys of twins. We used our model to predict views and diagnoses for 10 sets of twins (n=20 fetuses) including TOF and HLHS. Sensitivity and specificity were 100% and 72% (**Table S1**).

Models should be robust to minor variation in image quality to be useful for a range of patients and medical centers. We therefore assessed model performance on images within OB-125 that expert clinicians did not label as high-quality views, but that the model did classify as target views (**Figure 2d, 2f**). We inspected these “false-positive” images directly and analyzed their prediction probabilities. Of images with probability ≥ 0.9, two thirds (66%) were in fact target views, but of lower quality (e.g. slightly off-axis, heavily shadowed) than ones chosen by experts, and most (59%) of these low-quality target views had probabilities ≥ 0.9 (**Figure S3**). Therefore, the model can appropriately detect target views of lower quality. We submitted these lower-quality target images for diagnostic prediction and found sensitivity of 95% (95% CI, 83-99%) and specificity of 39% (95% CI, 28-50%). Thus, the ensemble model can make use of sub-optimal images in fetal surveys to detect complex CHD, albeit with lower specificity.

As with view classification above, we performed several analyses to determine whether the diagnostic classifications were based on clinically relevant image features. We trained a set of per-view binary classifiers for each of the two most common lesions in our dataset—TOF and HLHS—and examined ROC curves, saliency maps, and Grad-CAMs. For TOF, AUCs were highest for the two views from which TOF is most easily clinically appreciable: 3VT and 3VV (**Figure 3b**). For HLHS, 3VT, 3VV, A5C, and A4C are all abnormal, consistent with higher AUCs in **Figure 3c**. Saliency mapping and Grad-CAM highlighted pixels and image regions relevant to distinguishing these lesions from normal (**Figure 3g**). In clinical practice, reported sensitivity in detecting TOF and HLHS is as low as 50 and 30%, respectively^37^. With our model, sensitivity is 71% for TOF and 89% for HLHS (specificity 89% and 92%; **Table S1**).

### Segmentation for fetal biometrics

Biometric measurements aid in fetal CHD screening and diagnosis^12^. We therefore trained a modified U-Net^28^ (**Figure S1b, Methods**) to find cardiothoracic structures in A4C images and used these segmented structures to calculate CTR, CA, and FAC for each cardiac chamber (**Table 2, Figure 4**). Normal, TOF, and HLHS hearts were represented in training and testing. Per-class Jaccard similarities measuring overlap of labeled and predicted segmentations are found in **Table S2**. Predictably, Jaccards were higher for more highly represented pixel classes (e.g., background) and were similar to intra-labeler Jaccards (range 0.53-0.98, mean 0.76). Example labels and predictions for segmented structures are shown in **Figure 4**.

**Table 2.**
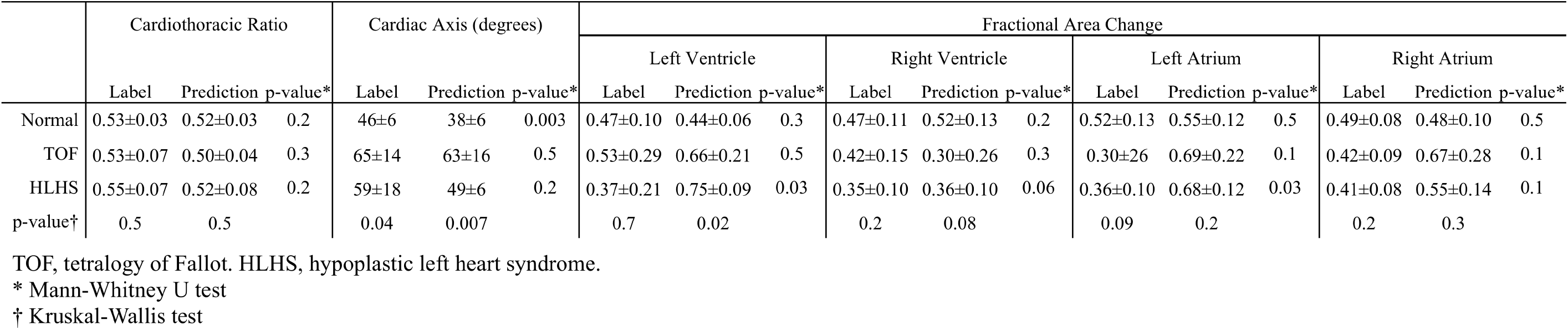
Fetal Biometrics calculated from pixel-level segmentation of anatomic structures.

**Figure 4.**
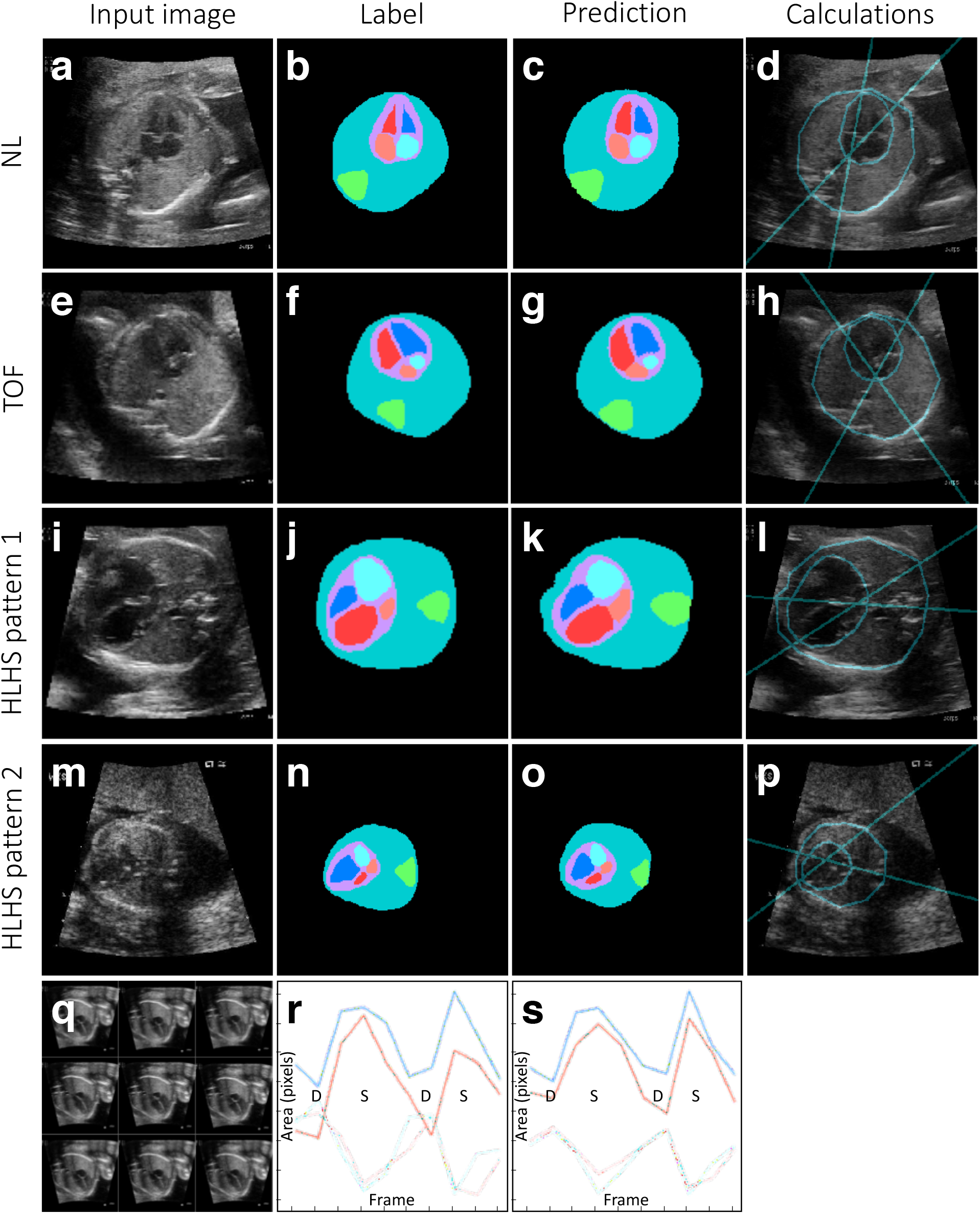
Fetal cardiac structure and function from segmentation. Example input image, ground truth label of anatomic structures, prediction of anatomic structures, and calculations of cardiothoracic ratio and cardiac axis for a normal heart (a-d), tetralogy of Fallot (TOF; e-h), and hypoplastic left heart syndrome (HLHS; i-p). Segmentation of an image series (q) allows plots of chamber area over time (label, r; prediction, s) and identification of image frames in ventricular systole (S) and diastole (D) for fractional area change calculation. Teal, thorax; green, spine; purple, heart; red, left ventricle; pink, left atrium; blue, right ventricle; light blue, right atrium.

Normal cardiothoracic circumference ratios range from 0.5-0.6^1^. Mann-Whitney U (MWU) testing showed no statistical differences among clinically measured and labeled CTR for normal hearts, nor between labeled and model-predicted CTR. CTR for TOF and HLHS hearts were normal, as previously reported^1^.

A normal cardiac axis is 45 ±20 degrees^12^. Consistent with the literature^38^, mean cardiac axis was increased in TOF at 63±16 degrees (range 54-80; p-value 0.007). CA for HLHS was not found in the literature, but model-predicted CA was 49±2 degrees (range 33-72; p-value 0.04).

In addition to the five still-image views, it is best practice to also obtain a video of the A4C view to assess cardiac function^1^. FAC quantifies this assessment. From a study measuring 70 normal 18-24 week fetuses, 50^th^ percentile for left and right ventricular FAC averaged 0.34±0.01 and 0.33±0.02, respectively^39^. In our test dataset, labeled FAC for normal LV and RV were 0.48±0.09, and model predictions were 0.47±0.10 (p-value 0.3), and 0.47±0.11 (p-value 0.2), respectively. Although there are no fetal atrial FAC values established in the literature, model-predicted LA and RA FAC were statistically indistinguishable from labels at 0.52±0.12 and 0.49±0.08, respectively (p-values 0.5 and 0.5). All measurements are summarized in **Table 2** and **Figure S2**.

Taken together, the data show that fetal cardiothoracic biometrics can be derived from image segmentation, showing good agreement between previously reported values and the potential to provide additional metrics not yet benchmarked.

## Discussion

With clear benefit to early diagnosis and treatment of CHD, and growing research on *in utero* interventions, the need for accurate, scalable fetal screening for CHD has never been stronger^40^, while sensitivity and specificity for CHD detection remain low at centers and clinics worldwide^1^. To address this, we investigated the impact of combining real-world fetal ultrasound and trusted clinical guidelines with cutting-edge deep learning to achieve expert-level CHD detection from fetal surveys, one of the most difficult diagnostic challenges in ultrasound. In over 4000 fetal surveys (over 4M images), the ensemble model achieved an AUC of 0.99.

Deep learning has been used on various medical tasks^21,23,41^, but to our knowledge, this is the first use of deep learning to approximately double community-level sensitivity and specificity on a global diagnostic challenge in a population-based test set.

The model’s performance and speed allow its integration into clinical practice as software onboard ultrasound machines to improve real-time acquisition and to facilitate telehealth approaches to prenatal care which are so sorely needed^9^. As a key benefit, the view classifier could be used on its own to help ensure adequate view acquisition. For retrospectively collected images, the model could be used as standalone software where a user uploads a study and receives model-chosen views and diagnostic predictions.

### Strengths of this study

#### Generalizability

To ensure our model could work robustly in real-world settings, we used two-dimensional ultrasound and standard recommended fetal views rather than rather than specialized or vendor-specific image acquisitions^42,43^. Furthermore, we tested our model in a range of different scenarios and on different independent test datasets. Importantly, the model maintained high sensitivity on external imaging, sub-optimal imaging, imaging from fetal surveys, from fetal echocardiograms, on datasets with community-level CHD prevalence, and with high CHD prevalence. Where a test dataset approximately 10% of the size of the training dataset has arisen as an informal rule of thumb for adequate testing in the data science community, we tested on over 350% of the number of studies in the training set, and over 4000% the number of images.

#### Interpretability

Our approach to both model design and testing ensured interpretability at several levels, which can help with clinical adoption. Choosing to use an ensemble of classifiers—first a view detector, then per-view diagnostic classifiers, and finally a classifier for composite diagnosis—allowed us to incorporate clinical view recommendations into our model and to demonstrate that model performance per view and per CHD lesion were consistent with clinical knowledge about which views were most likely to aid in detection of specific lesions.

Analysis of confusion matrices, ROC curves, and incorrectly classified images helped determine that model error mirrored uncertainties in clinical practice. Saliency mapping and Grad-CAM for both view and diagnostic classifications demonstrated that model predictions relied on cardiac structures. The prominence of the aorta, the right heart, and the stomach as distinguishing features among the five target views is both novel and sensible. A comparison of the different testing scenarios (**Table S1**) suggests that both the quality of images and the number of available images per study contribute to the best overall performance.

#### Novel approaches to training

As mentioned above, we incorporated two similar study types— fetal echocardiograms and fetal surveys—in a multi-modal approach to model training that harnessed more specialized imaging in service of improving performance on screening imaging. By feeding only target views into the diagnostic classifier step, we took a more data-efficient approach to the diagnostic classifier compared to using the entire ultrasound. We also took a novel approach to addressing variation in image quality that relied on human experts to agree only on labeling diagnostic-quality images for training (in testing, the model analyzed all images). This approach economized on human capital, consolidating inter-expert agreement on diagnostic-quality images, while providing fewer constraints to the model training, since some aspects that make an image low-quality to a human eye may not matter as much to a computer “eye” (image contrast is a good example of this). We found that prediction probability was an indirect representation of the model’s quality assessment, and that using cutoffs for high-prediction-probability images improved model performance.

#### Diagnostic signals from small/lean datasets and rare diseases

While it is the most common birth defect, CHD is still relatively rare. Moreover, unlike modalities like photographs^21,23^, ECG^41^ or chest X-ray, each ultrasound study contains thousands of image frames. Therefore, designing a model that could work on a large number of non-independent images from a relatively small subject dataset was an important challenge to overcome. Taken together, the strengths above allowed us to find diagnostic signals for rare diseases and allowed computational efficiency both in training and in subsequent predictions on new data, which is key to translating this work toward real-world and resource-poor settings where it is needed^44^.

### Limitations of this study

While 4,108 fetal surveys a significant test set especially when considering the size of each ultrasound, hundreds of millions of fetal surveys are performed annually at many thousands of medical centers and clinics worldwide. Therefore, expanded testing of the model prospectively and in multiple centers, including community/non-expert centers, will be important going forward. It will also be important to test the model on imaging that includes a range of non-cardiac malformations. Several small improvements in model algorithms, as well as more training data from more centers, may further boost performance and may allow for diagnosis of specific lesion types. Similarly, more training data for image segmentation, including segmenting additional CHD lesions, will improve segmentation model performance and allow those results to be integrated into the composite diagnostic classifier. Further clinical validation of segmentation-derived fetal biometrics will be needed, particularly where metrics on particular CHD lesions have not yet been described elsewhere.

We look forward to testing and refining ensemble learning models in larger populations in an effort to democratize the expertise of fetal cardiology experts to providers and patients worldwide, and to applying similar techniques to other diagnostic challenges in medical imaging.

## Data Availability

Due to the sensitive nature of patient data (and especially fetuses as a vulnerable population), we are not able to make these data publicly available at this time.

## Acknowledgements

We thank Drs. Atul Butte, Deepak Srivastava, and Ramy Arnaout for critical reading of the manuscript; Drs. Michael Brook, Marc Kohli, Wayne Tworetzky, and Kathy Jenkins for facilitating data access. We thank all clinicians who served as human subjects, including Christine Springston RDCS, and Drs. Katherine Kosiv, Christiana Tai and David Abel; others wished to remain anonymous. This project was also supported by the UCSF Academic Research Systems and the National Center for Advancing Translational Sciences, National Institutes of Health, through UCSF-CTSI Grant UL1 TR991872.

## Author Contributions

R.A. and A.M. conceived of the study. R.A. and E.C. designed and implemented all computational aspects of image processing, data labeling, pipeline design, neural network design, tuning, and testing, and data visualizations. R.A., L.C., Y.Z., and A.M. labeled and validated images. J.C.L. curated and sent external data. R.A. wrote the manuscript with critical input from A.M., E.C., and all authors.

## Competing Interests

Some methods used in this work have been filed in a provisional patent application.

## Supplemental Tables

**Table S1.**
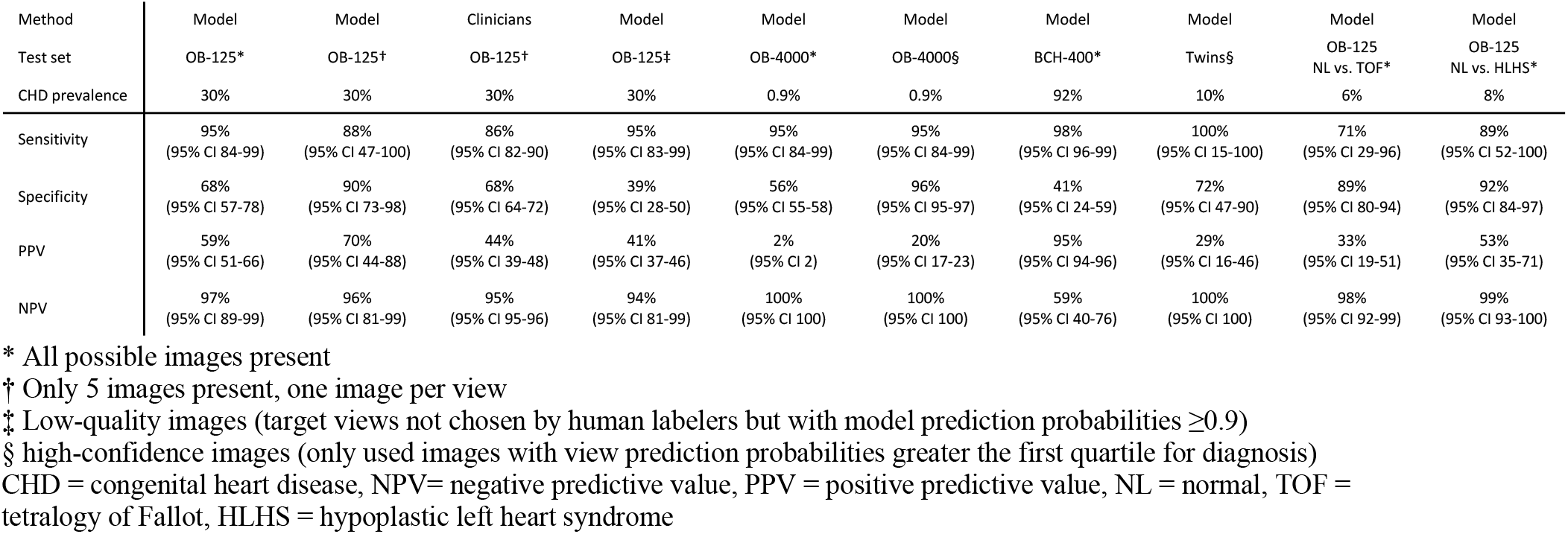
Summary of diagnostic performance in different test cases. Test threshold chosen from OB-4000§ ROC curve (Figure 3e) to optimize sensitivity. CHD prevalence is again shown to aid in interpretation of predictive values.

**Table S2.** Average Jaccard similarities for labeled and predicted anatomic structures.

| Structure | thorax | heart | RA | RV | LA | LV | spine | bkgnd |
| --- | --- | --- | --- | --- | --- | --- | --- | --- |
| Overall | 0.79 | 0.86 | 0.77 | 0.70 | 0.63 | 0.60 | 0.67 | 0.99 |
| Normal | 0.80 | 0.87 | 0.82 | 0.77 | 0.72 | 0.78 | 0.69 | 0.99 |
| TOF | 0.73 | 0.86 | 0.81 | 0.63 | 0.64 | 0.65 | 0.47 | 0.98 |
| HLHS | 0.77 | 0.82 | 0.66 | 0.59 | 0.47 | 0.29 | 0.62 | 0.99 |

TOF, tetralogy of Fallot; HLHS, hypoplastic left heart syndrome; RA, right atrium;
RV, right ventricle; LA, left atrium; LV, left ventricle.

## Supplemental Figures Legends and Figures

**Figure S1.**
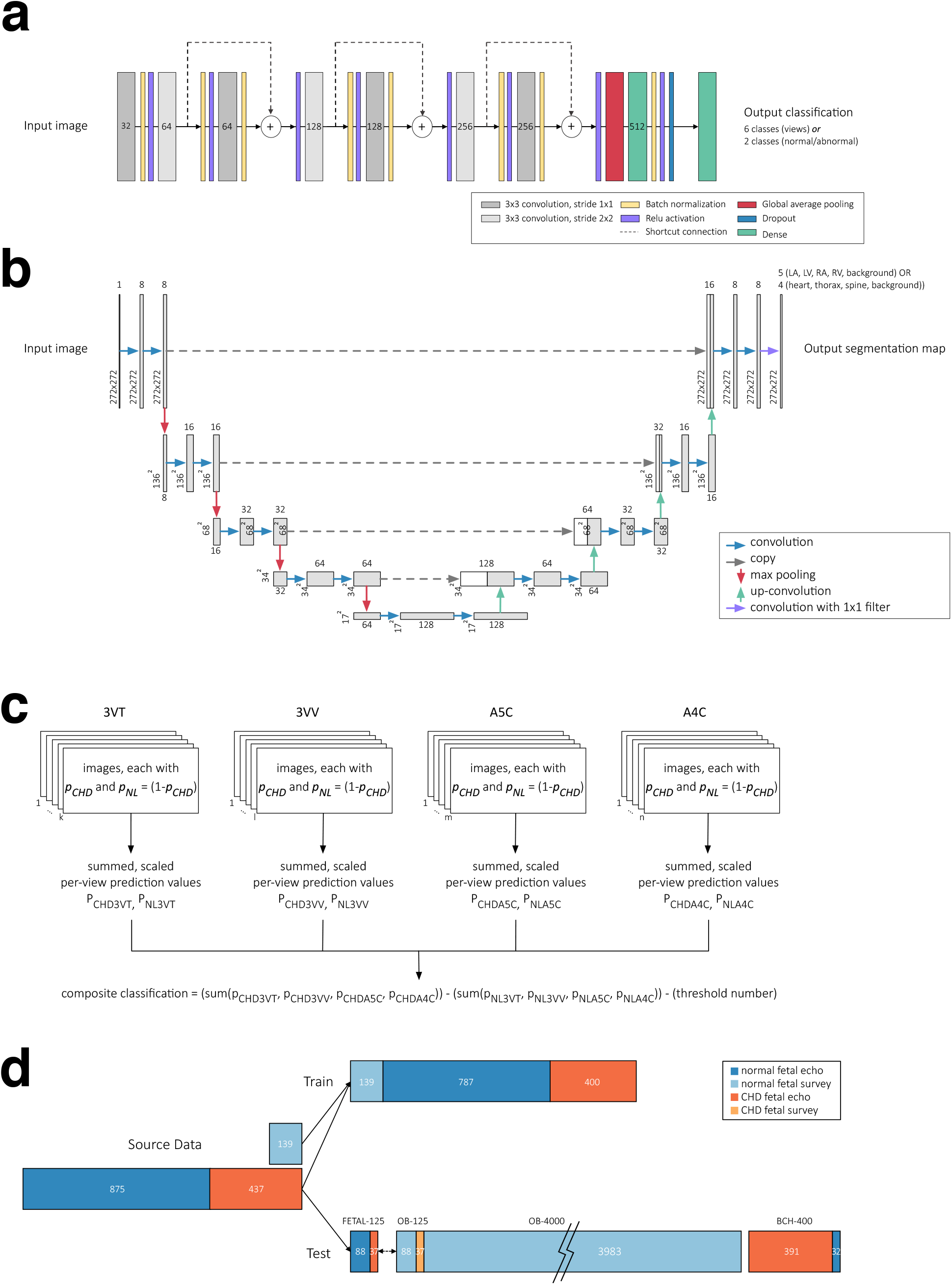
Neural network architectures, schematic of rules-based classifier, and schematic of training and test data. (a) Neural network architecture used for classification, based on ResNet (He et. al. 2015). Numbers indicate the number of filters in each layer, while the legend indicates the type of layer. For convolutional layers (grey), the size and stride of the convolutional filters is indicated in the legend. (b) Neural network architecture used for segmentation, based on UNet (Ronneberger et. al. 2015). Numbers indicate the pixel dimensions at each layer. (c) A schematic for the rules-based classifier (“Composite dx classifier,” Figure 1b) used to unite per-view, per-image predictions from neural network classifiers into a composite (per-heart) prediction of normal vs. CHD. Only views with AUC > 0.85 (Figure 3a) were used. For each view, there are various numbers of images k,l,m,n, each with a per-image prediction probability *p*_*CHD*_ *and p*_*NL*_. For each view, per-image *p*_*CHD*_ *and p*_*NL*_ were summed and scaled (see Methods) into a pair of overall prediction values for each view (e.g. P_CHD3VT_ and P_NL3VT_). These are in turn summed for a composite classification. Evaluating true positive, false positive, true negative, and false negative with different thresholds allowed construction of a ROC curve (Figure 3e). 3VT, 3-vessel trachea. 3VV, 3-vessel view. A5C, apical 5-chamber. A4C, apical 4-chamber. (d) A graphical depiction of the five UCSF and BCH training and test datasets. Dotted line between FETAL-125 and OB-125 indicates that these are corresponding imaging from the same fetuses. OB-4000 includes OB-125. Broken lines in OB-4000 indicate it is too large to be drawn to scale. Normal fetal echocardiograms, dark blue; normal fetal surveys, light blue; CHD fetal echocardiograms, dark orange; CHD fetal surveys, light orange.

**Figure S2.**
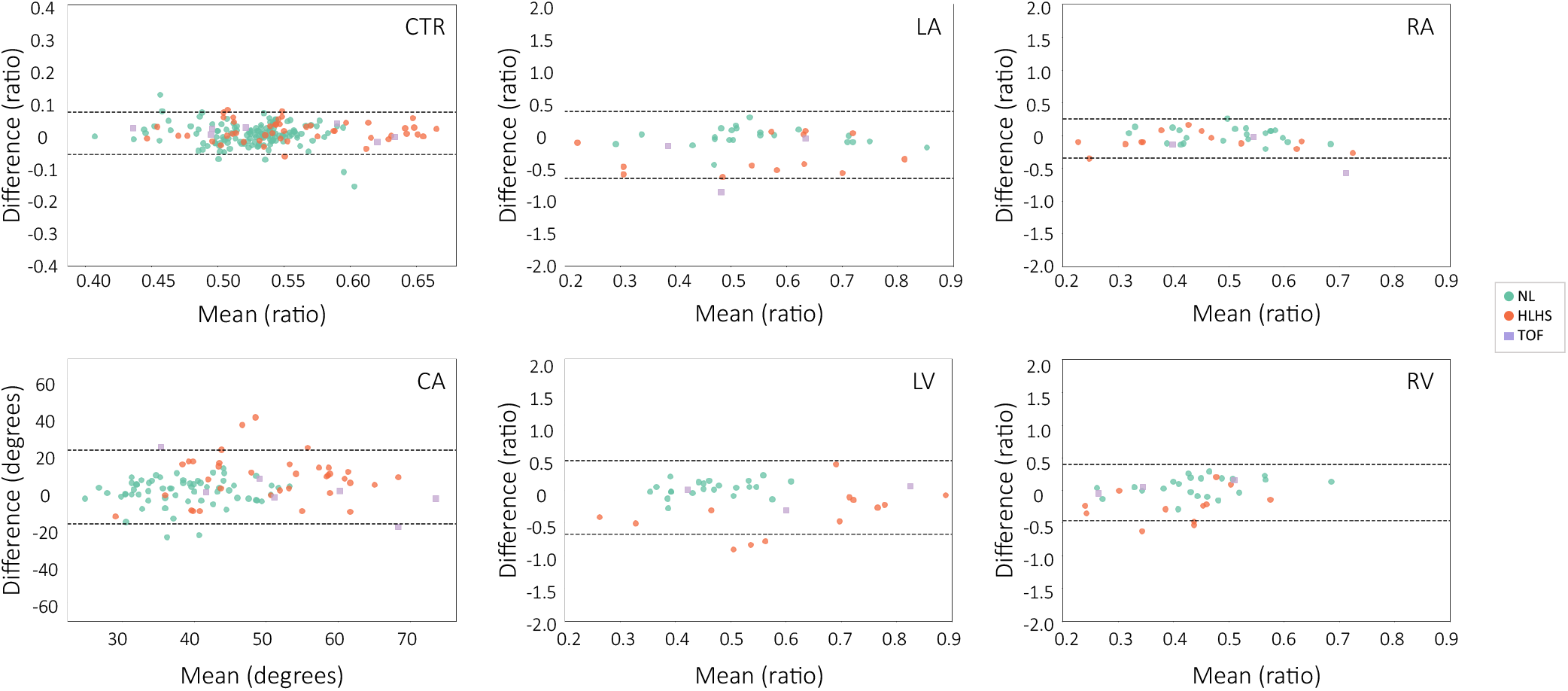
Bland-Altman plots comparing cardiac measurements from labeled vs. predicted structures. CTR, cardiothoracic ratio; CA, cardiac axis; LV, left ventricle; RV, right ventricle; LA, left atrium, RA, right atrium. Legend indicates measures for normal hearts (NL), hypoplastic left heart syndrome (HLHS), and tetralogy of Fallot (TOF).

**Figure S3.**
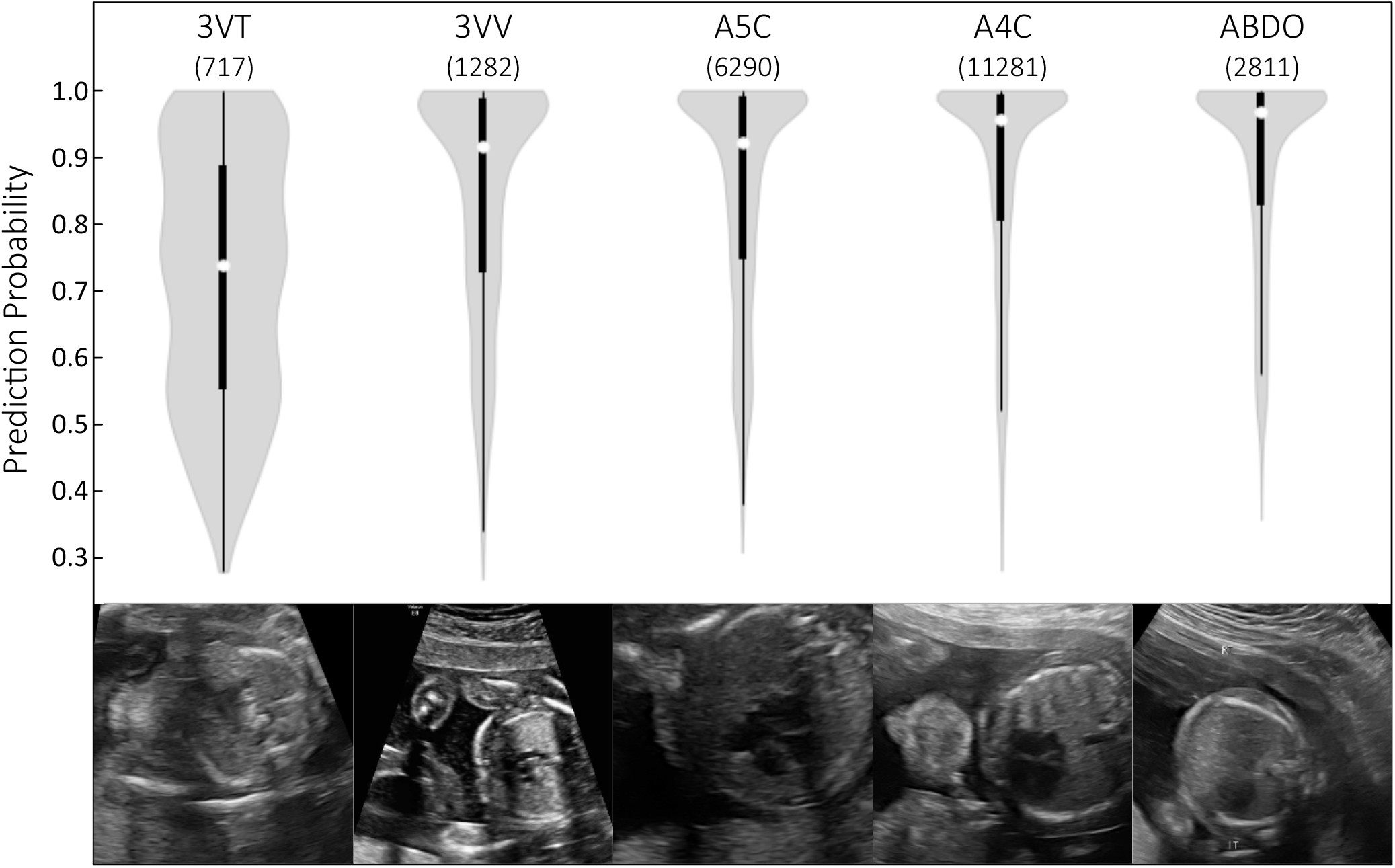
Model confidence on sub-optimal images. Examples of sub-optimal quality images (target views found by the model but deemed low-quality by human experts) are shown for each view, along with violin plots showing prediction probabilities assigned to the sub-optimal target images (White dots signify mean, thick black line signifies 1^st^ to 3^rd^ quartiles).

## Notes

### Funding Statement

No entity other than the authors listed played any role in the design of the study; the collection, analysis, or interpretation of data; writing of the report; or in the decision to submit the paper for publication. This work was supported by the American Heart Association, the National Institutes of Health, and the Department of Defense.

### Author Declarations

UCSF Boston Children's Hospital

